# Simulated resections and RNS placement can optimize post-operative seizure outcomes when guided by fast ripple networks

**DOI:** 10.1101/2024.03.26.24304802

**Authors:** Shennan Aibel Weiss, Michael R. Sperling, Jerome Engel, Anli Liu, Itzhak Fried, Chengyuan Wu, Werner Doyle, Charles Mikell, Sima Mofakham, Noriko Salamon, Myung Shin Sim, Anatol Bragin, Richard Staba

## Abstract

In medication-resistant epilepsy, the goal of epilepsy surgery is to make a patient seizure free with a resection/ablation that is as small as possible to minimize morbidity. The standard of care in planning the margins of epilepsy surgery involves electroclinical delineation of the seizure onset zone (SOZ) and incorporation of neuroimaging findings from MRI, PET, SPECT, and MEG modalities. Resecting cortical tissue generating high-frequency oscillations (HFOs) has been investigated as a more efficacious alternative to targeting the SOZ. In this study, we used a support vector machine (SVM), with four distinct fast ripple (FR: 350-600 Hz on oscillations, 200-600 Hz on spikes) metrics as factors. These metrics included the FR resection ratio (RR), a spatial FR network measure, and two temporal FR network measures. The SVM was trained by the value of these four factors with respect to the actual resection boundaries and actual seizure free labels of 18 patients with medically refractory focal epilepsy. Leave one out cross-validation of the trained SVM in this training set had an accuracy of 0.78. We next used a simulated iterative virtual resection targeting the FR sites that were highest rate and showed most temporal autonomy. The trained SVM utilized the four virtual FR metrics to predict virtual seizure freedom. In all but one of the nine patients seizure free after surgery, we found that the virtual resections sufficient for virtual seizure freedom were larger in volume (p<0.05). In nine patients who were not seizure free, a larger virtual resection made five virtually seizure free. We also examined 10 medically refractory focal epilepsy patients implanted with the responsive neurostimulator system (RNS) and virtually targeted the RNS stimulation contacts proximal to sites generating FR at highest rates to determine if the simulated value of the stimulated SOZ and stimulated FR metrics would trend toward those patients with a better seizure outcome. Our results suggest: 1) FR measures can accurately predict whether a resection, defined by the standard of care, will result in seizure freedom; 2) utilizing FR alone for planning an efficacious surgery can be associated with larger resections; 3) when FR metrics predict the standard of care resection will fail, amending the boundaries of the planned resection with certain FR generating sites may improve outcome; and 4) more work is required to determine if targeting RNS stimulation contact proximal to FR generating sites will improve seizure outcome.

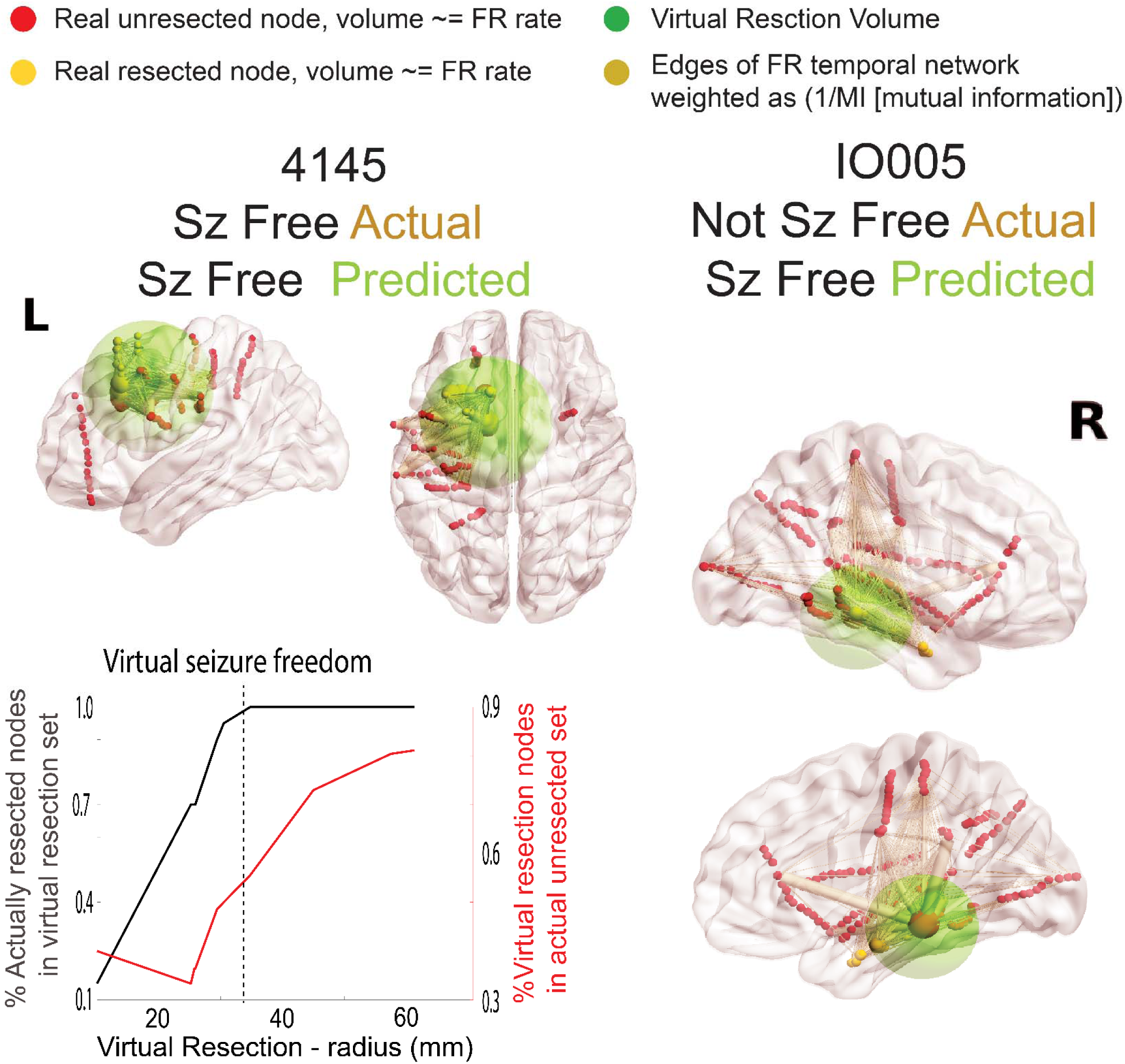
Graphical Abstract.

## Introduction

The standard of care for planning an efficacious epilepsy surgery, with minimal morbidity, is electroclinical delineation of the seizure onset zone (SOZ) in the epilepsy monitoring unit by an expert epileptologist, integrated with neuroimaging findings from MRI, PET, SPECT, and MEG modalities^1^ and consideration of seizure semiology. Even if the SOZ is sufficiently sampled by the stereo EEG (SEEG) implant, resection of the SOZ does not always correlate with seizure outcome^2–7^, leaving a considerable percentage of surgical patients with uncontrolled seizures^8–10^. This is especially true for patients with non-lesional frontal lobe epilepsy^8^. However, utilizing the rates (events/min) of interictal high-frequency oscillations (HFOs: 80-600 Hz) could be an alternative to the clinical standard of care of delineating the boundaries of the SOZ^11,12^.

Many studies have shown HFO and HFO related biomarkers such as spike-ripples^13–15^, the ripple-spike cross rate^16^, and entropy measures^17^, computed from inter-ictal epochs during non-REM sleep, strongly correlate the SOZ or resected territory in seizure free patients. In the current study, we focused on a subpopulation of HFOs known as fast ripples (200-600 Hz, FR), which are brief (8-50 ms) bursts of oscillatory activity that are, in most context, pathological^18–21^. In retrospective studies, resecting 60% of FR (i.e., FR resection ratio [RR]) had a 70-80% accuracy for predicting seizure freedom^12^, concluding the cortical territory generating FR are necessary and sufficient for seizure generation^2,11,12,22–28^ and thereby demarcate the epileptogenic zone^1^. This hypothesis is problematic because microelectrode studies show FR can occur at high rates contralateral to the SOZ^23,24,29^, and similar findings have been reported in murine models of epileptogenesis^30,31^. Also, since seizure free outcomes can be achieved with only a 60% FR resection ratio (*i.e.,* 40% of FR left intact) it suggests that certain cortical FR sites are more important than others for seizure generation, even if all FR are pathological per se.

In contrast to the epileptogenic zone^1^, other epilepsy researchers have conceptualized that an epileptic network is responsible for seizure generation^10,32,33^. The epileptic network can be formulated in diverse ways and with many substrates. We have proposed that FR are one important substrate of the epileptic network because: 1) FR propagate primarily within the SOZ^34,35^; 2) propagating FR, and FR with increased excitability, can prime epileptiform spike discharge^34,36^; 3) prior to seizure onset larger amplitude FR superimposed on pre-ictal spikes may be trigger the seizure^37–39^; and 4) surgically targeting autonomous, high-rate cortical FR sites^40,41^ are important for a seizure free outcome^42,43^. Based on these findings we derived metrics using graph theoretical analysis of spatial networks and FR temporal correlations. The spatial FR graph theoretic metric does not have a true anatomic correlation but overcomes spatial sampling bias inherent in the FR RR^43^. The temporal FR graph metrics are neurophysiologically relevant as they correspond to the synchrony of FR across all the sampled FR sites or nodes^43^.

In the current study, we detected HFOs in SEEG recordings during non-REM (NREM) sleep from 18 patients, and derived the FR RR, the spatial FR network measure, and two temporal FR network measures based on the actual resection or ablation. We used machine learning to test if using these four metrics together could classify post-operative outcome using leave one out cross-validation. The trained machine was then tested using virtual resections that targeted autonomous, high rate cortical FR sites. We found in 8 of 9 patients who were seizure free after resection, the virtual resection was anatomically larger. In five patients who were not seizure free after surgery, amending cortical regions from the virtual resection predicted a seizure free outcome. Lastly, in 10 patients who had a responsive neurostimulator system (RNS), we simulated changes in the location of the RNS stimulation contacts and in several subjects, targeting electrical stimulation to high-rate FR sites suggested the seizure outcome may improve from intermediate to super-responders (>90% seizure reduction).

## Methods

### Patients

Consecutive recordings selected from 19 patients who underwent intracranial monitoring with depth electrodes between 2014 and 2018 at the University of California Los Angeles (UCLA) and from 29 patients at the Thomas Jefferson University (TJU) in 2016–2018 for the purpose of localization of the SOZ. Data collection was planned before the study was conceptualized. Among these 48 patients, 31 underwent resections and ablations and 12 were implanted with RNS. Inclusion criteria for this study included pre-surgical MRI for MRI-guided stereotactic electrode implantation, as well as a post-implant CT scan to localize the electrodes, and stereo EEG recordings during non-rapid eye movement (REM) sleep at a 2 kHz sampling rate. Patients were excluded if: 1) no resection/ablation or RNS placement was performed; 2) a post-resection/ablation MRI or a post-RNS implant CT was not obtained; 3) no adequate post-operative clinical follow up; 4) A failure to record at least ten minutes of artifact free iEEG during non-REM sleep; and 5) graph theoretical analysis indicated incomplete or poor spatial sampling^43^. Based on these criteria 18 patients were included in the analysis of resection/ablation outcome, and 10 patients in the analysis of RNS outcome (Figure 1). All patients gave verbal and written informed consent prior to participating in this research, which was approved by the University of California Los Angeles and Thomas Jefferson University institutional review boards (IRBs). Eligible patients were found through queries of pre-existing clinical databases. The methods in this paper adhered, and were in accord with, the relevant guidelines and regulations of the IRBs.

**Figure 1:**
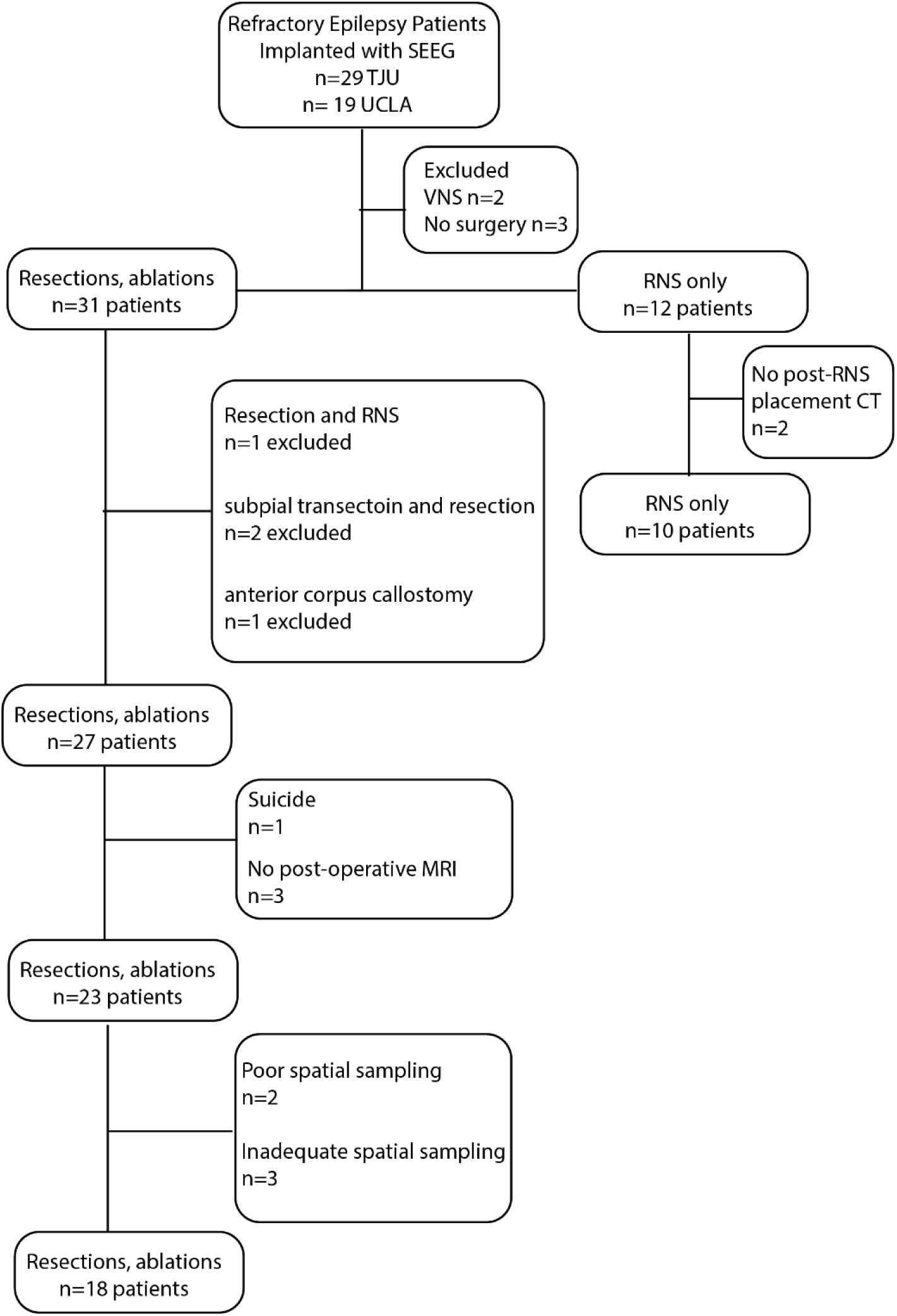
Flow chart of patient enrollment with inclusion and exclusion criteria.

### Neuroimaging

Using an in-house pipeline (https://github.com/pennmem/neurorad_pipeline), T1-pre-implant and post-resection MRIs were obtained for each patient. Post-implantation SEEG and RNS CT scans were then co-registered and normalized with the MRIs using Advanced Neuroimaging Tools (ANTs)^44^ with neuroradiologist supervision. The position of each electrode contact in the post-SEEG implant CT and post-RNS placement CT was localized to normalized MNI coordinates and the Desikan-Killiany atlas^45^. Identification of the named electrode contacts in the resection cavity was performed manually in itk-SNAP.

### EEG recordings and HFO detection

Clinical iEEG (0.1–600 Hz; 2000 samples per second) was recorded from 8 to 16 depth electrodes, each with 7–15 contacts, using a Nihon-Kohden 256-channel JE-120 long-term monitoring system (Nihon-Kohden America, Foothill Ranch, CA, USA), for each patient. A larger number of electrodes with more contacts were implanted at TJU. For the recordings performed at UCLA. the reference signal used for was a scalp electrode position at Fz. The reference signal used for the TJU recordings was an electrode in the white matter. One to two days after implantation, for each patient a 10–60 min iEEG recording from all the depth electrodes that contained large amplitude, delta-frequency slow waves (i.e., non-REM sleep) was selected for analysis. Only iEEG that was free of low levels of muscle contamination and other artefacts was selected. HFOs, HFOs superimposed on spikes and sharp-spikes were detected in the non-REM sleep iEEG using previously published methods (https://github.com/shenweiss)^46–50^ implemented in MATLAB (Mathworks, Natick, MA, USA)^43^. Identification and quantification of HFO on spikes was performed by the topographical analysis of the wavelet convolution^46^. Following automatic detection of HFO and sharp-spikes, false detections of clear muscle and mechanical artifact were removed by visual review in Micromed Brainquick (Venice, Italy). The seizure onset zone was clinically delineated and aggregated during the entirety of the epilepsy monitoring unit evaluation.

### Calculation of resection ratios

The seizure onset zone (SOZ) resection ratio was calculated as the number of resected SOZ contacts divided by the total number of SOZ contacts. The RRs for fast ripple (FR) >350 Hz [*i.e.,* fast ripple on oscillation > 350 Hz, and all fast ripple on spike]^42,43,51^, all FR (200-600 Hz), ripple on spike (RonS), and ripple on oscillation (RonO) were calculated as the number of total number of the events recorded from resected electrode contacts in the numerator, and the total number of the events recorded by all the electrode contacts in the denominator. The RR values were calculated at each iteration of the simulated virtual resections (see: Virtual Resections and Outcome Prediction).

### Derivation of FR graph theoretical measurements

All graph theoretical measures were calculated using the Brain Connectivity Toolbox (https://sites.google.com/site/bctnet/)^52^. The weighted edges for the spatial FR net (FR rate–distance radius resected difference *i.e.,* RDRRD) was calculated by the average rate (/min) of the FR>350 Hz [*i.e.,* fast ripples on oscillations>350 Hz, and all fast ripple on spikes] recorded by two respective nodes multiplied by the Euclidian distance (mm) between these nodes. One graph used all sampled FR>350 Hz generating nodes and another graph only the resected FR>350 Hz generating nodes. The spatial FR net was defined as the square root of the difference between the radius of the whole brain graph and the radius of the resected only brain graph. The radius of the actual resection was computed as the radius of the graph of the weighted graph with edges defined by the Euclidian distance between resected nodes alone. The edges for the mutual information (MI) networks were calculated using FR>350 Hz event ‘spike trains’ defined by the onset times of each event and then calculating MI between nodes using the adaptive partition using inter-spike intervals MI estimator.^53^ Using these adjacency matrices, and their inverses, the temporal FR net-A (gammaRR) and B (urmLE) were calculated^43^. In brief temporal FR net-A was defined as the path length computed from the resected nodes alone as the numerator and the path length of the whole network in the denominator. Temporal FR net-B was defined as the mean nodal local efficiency (LE) across all nodes with a LE greater than zero. In each iteration of the virtual resection the set of virtual resected nodes and virtual unresected nodes were used to derive virtual values for spatial FR net, and temporal FR net-A, B (see: Virtual Resections and Outcome Prediction).

### Machine learning using a support vector machine (SVM)

Support vector machines (SVM) were trained using the dichotomized labels of seizure free and non-seizure free for each patient’s actual outcome and the factors: 1) FR RR; 2) spatial FRnet; 3) temporal FRnet-A; and 4) temporal FRnet-B derived from FR>350 Hz and the actual resection/ablation boundaries. The SVM was trained after normalizing the data and using a Radial Basis Function kernel that is automatically scaled to reduce the effect of outliers on SVM training^42^. Gamma was calculated as 1/number of factors, and C was defined as 1. Following SVM training, leave one out cross-validation was performed with 18 folds, and accuracy was interpreted as 1 minus k-fold loss. The SVM was then tested on the virtual values of 1) FR RR; 2) spatial FRnet; 3) temporal FRnet-A; and 4) temporal FRnet-B from each iteration of the virtual resections in the 18 patients to label virtual seizure freedom (see: Virtual Resections and Outcome Prediction). We selected an SVM for classification rather than a mixed regression models because the data was not assumed to be in a hierarchical structure, and we were not interested in describing the random effects. We also selected a SVM instead of a multiple regression model due to the focus on a dichotomized outcome (*i.e.,* seizure free or not) and assumed non-linearity of the four factors.

### Virtual Resections and Outcome Prediction

The first virtual resection volume was determined by defining all the graph nodes (i.e., contacts) with a FR>350 Hz rate > 1/minute as a candidate set and finding the node with the smallest LE as the candidate node in the candidate set. If no contacts had a FR rate > 1 minute, or no such nodes remained in the candidate set, all nodes were included in the candidate set and the candidate node was selected as the node with the highest FR rate. The candidate node served as the center of the sphere of the virtual resection(s). A resection sphere with a 1 cm radius was initially simulated, centered on this first candidate node, and all nodes falling within this sphere were included in the virtual resected set after excluding contralateral contacts. For all the nodes in the virtual resected set and unresected set, the virtual SOZ RR, RonS RR, FR RR, spatial FRnet, and temporal FRnet-A, B were calculated. Additionally, we quantified the proportion of overlapping and non-overlapping nodes in the virtual resected set and the set of nodes in the actual resection. Then, in the second iteration of the simulation, the node with second lowest LE, or second highest FR rate, was included in the resected set. The radius between the first node and this second resected node, with an additional 1 cm buffer, was used to calculate a second sphere and define the new resected set. Iteration of the simulation continued through all the candidate nodes in the candidate set with an incrementally increasing, but not decreasing, radius. For each iteration of the simulation, the SVM predicted whether the virtual outcome was seizure free. Areas of the brain that were not sampled by SEEG contacts, including outside of the brain, did not influence the FR metrics or the SVM label. The 1 cm margin around the node of interest used by the simulations was selected per our Neurosurgical collaborators’ expertise (Figure 5). If the radius extended into three brain lobes, then the virtual resection simulation was stopped and the outcome was designated as non-seizure free.

### Virtual RNS stimulation lead placement and RNS metrics

We examined 10 patients implanted with the responsive neurostimulation (RNS) device and asked if alternate placement of the RNS stimulation contacts at sites generating FR>350 Hz at high rates would predict a better seizure outcome. To approximate the brain regions that were maximally stimulated by the actual and virtual RNS placements, we defined the pre-implant SEEG electrode stimulated contacts as within a radius of <1.5 cm of the eight RNS contacts (i.e., two leads of either a four-contact depth or subdural strip)^54^. Our calculations were based on the magnitude of the electrical field generated by monopolar current sources of 1-3 mAmp ^54–57^. We calculated the SOZ stimulation ratio (SR), FR SR, and the FR stimulated global efficiency (SGe), herein described as RNS temporal FR net^59^ using the boundaries of the calculated stimulated brain regions. The RNS temporal FR net was derived by calculating the efficiency using an adjacency matrix of the mutual information (MI) between FR spike trains, defined by the FR onset time, between stimulated and first-degree neighboring contact pairs. We then asked if these values differed in patients with a super responder (>90% seizure reduction) clinical outcome^59^. Then, the 10 patients with RNS placement were subdivided into those with bilateral and unilateral placement of the RNS stimulation leads. Virtual RNS stimulation contacts were selected contiguous to the pre-surgical stereo EEG contacts with highest FR rate. For patients with bilateral placement, we defined two sets of nodes, for each hemisphere, with the highest FR rate. We then calculated the corresponding virtual SOZ stimulation ratio, FR stimulation rate, and RNS temporal FRnet^54^ for each patient after virtual placement of the RNS stimulation contacts.

### Statistics

Values are expressed as mean +/− standard error of the mean (s.e.m). The Kruskall-Wallis test and Wilcoxon signrank test were implemented in MATLAB. Metrics of the contingency tables comparing virtual resections with actual resections (TP: true positive; TN: true negative; FP: false positive; FN: false negative) were calculated as: a) sensitivity= TP / (TP + FN); b) Specificity = TN / (TN + FP); c) positive predictive value [PPV] = TP / (TP + FP); d) negative predictive value [NPV] = TN / (TN + FN); e) Accuracy= (TP + TN) / (TP + TN + FP + FN); f) F1 Score = 2 * (PPV * Sensitivity) / (PPV + Sensitivity).

## Results

### Patient characteristics and spatial sampling limitations

After applying the exclusion criteria, the study cohort for patients that underwent resection consisted of 18 patients (7 males and 11 females) between the ages of 18 and 55 years old (Figure 1). These 18 patients had diverse epilepsy etiologies, including 4 with normal MRI findings^43^ and another 4 who had prior epilepsy surgery (Table 1). The neuroanatomic locations of the resections and ablations in this cohort were also diverse (Table 1). Post-operative seizure outcome was assessed 18 months or longer after surgery, except for one patient who died of sudden unexpected death in epilepsy six months after surgery (Table 1).

**Table 1:** Patient characteristics in the study cohort. Abbreviations L: left, R: right, N/A: not applicable, ATL: anterior temporal lobectomy, MTL: mesial temporal lobe, MTS: mesial temporal sclerosis, SMA: supplementary motor area, TBI: traumatic brain injury, LOC: loss of consciousness, RNS: responsive neurostimulator, VNS: vagal nerve stimulator, SUDEP: sudden unexpected death in epilepsy, @: time to last follow up. Rows colored blue indicate patients with poor spatial sampling (no electrodes recording FR on spikes at a rate > 1 minute), and those colored yellow as patients with incomplete spatial sampling (entire FR generating network resected despite non-seizure free outcome [see methods]). The number of nodes in the FR MI network with a local efficiency > 0 are listed with the patient ID in the first column.

| ID<br>#FR<br>MI nodes | Risk<br>Factor | MRI | PET<br>(hypo-<br>metabolic) | iEEG clinical<br>consensus SOZs | Surgery | Path. | Outcome |
| --- | --- | --- | --- | --- | --- | --- | --- |
| 1)<br>IO01<br>0 nodes<br>FR on<br>spike<br>positive | minor TBI | Normal | L temporal | L MT | modified L<br>ATL | Gliosis | Engel IA@24<br>months |
| 2)<br>IO08<br>27 Nodes | hyperten-<br>sive<br>enceph-<br>alopathy | post L ATL | N/A | L middle temporal<br>gyrus | modified L<br>temporal<br>lobectomy | Gliosis | Engel 1A @48<br>months |
| 3)<br>IO18<br>22 Nodes | Minor TBI | Normal | Normal | Right insula,<br>cuneus, inferior<br>and middle<br>frontal gyrus | R. Frontal lobe | Gliosis | Engel<br>IA@24<br>months |
| 4)<br>4122 | None | Normal | R<br>temporal | R Inferior temporal<br>gyrus | modified R<br>temporal<br>lobectomy | Gliosis | Engel IA@24<br>months |
| 5)<br>4124<br>4 Nodes | None | L MTL<br>white matter<br>hyperintensity | Normal | R SMA | R frontal lobe<br>resection | cortical<br>dysplasi<br>a | Engel IA@24<br>months |
| 6)<br>4145<br>32 Nodes | None | Normal | Normal | L cingulate gyrus,<br>medial frontal<br>gyrus, middle<br>frontal gyrus,<br>superior frontal<br>gyrus | L frontal lobectomy | cortical<br>dysplasi<br>a | Engel IA@40<br>months |
| 7)<br>4166<br>5 Nodes | meningitis | Encephalomala<br>cia | L<br>temporal | L MT, uncus,<br>superior<br>temporal gyrus,<br>frontal lesion | L temporal and<br>frontal lobe<br>resection | gliosis | Engel IB@42<br>months |
| 8)<br>IO12<br>9 Nodes | none | 1 cm pineal cyst | R lateral<br>temporal | L MT | Modified L<br>temporal<br>lobectomy | Gliosis | Engel<br>IIB@24<br>months |
| 9)<br>IO05<br>41 Nodes | febrile<br>seizures | prior<br>hippocampal<br>sparing<br>temporal<br>lobectomy | N/A | R anterior<br>cingulate, MT,<br>uncus | R ATL | gliosis | Engel<br>IVB@40<br>months |
| 10)<br>453<br>5 Nodes | None | T2 hyper-<br>intensity in R<br>temporal pole > L<br>frontal pole.<br>Inferior part of R<br>temporal pole<br>with blurred gray-<br>white matter<br>border | R<br>temporal | R MT | R anterior ATL | Cortical<br>dysplasia<br>IIb | Engel IA@60<br>months |
| 11)<br>456 | None | Normal | R<br>temporal | Bilateral MT,<br>middle temporal<br>gyrus R>L | modified R<br>ATL | gliosis | Engel<br>IVC@48<br>months |
| 12)<br>462<br>11 nodes | TBI,<br>family<br>history | left superior<br>temporal gyrus<br>encephalomalacia | L<br>parieto-<br>occipital | L temporal<br>neocortical, L<br>frontal | modified<br>LATL<br>hippocampus<br>sparing | gliosis | Engel IV@6<br>months RNS<br>placed and<br>revised |
| 13)<br>466<br>8 nodes | None | Normal | L<br>temporal | R fusiform gyrus,<br>superior temporal<br>gyrus, uncus | R ATL | MTS | Engel IB@35<br>months |
| 14)<br>473 | TBI w/<br>LOC | L MTS, extra-<br>temporal T2 | L<br>temporal<br>and<br>frontal | L MT, fusiform<br>gyrus, uncus | L MT Visualase | N/A | Engel IIIA@18<br>months |
| 15)<br>477<br>10 nodes | None | periventricular<br>nodular<br>heterotopia, right<br>frontal T2 | R<br>temporal | R MT | ATL | gliosis | Engel<br>IB@31<br>months |
| 16)<br>479 | TBI w/<br>LOC | Encephalomalacia | R<br>temporal | R insula, bi- lateral<br>middle temporal<br>gyrus, superior<br>temporal gyrus | modified R<br>ATL | gliosis | Engel<br>IVB@33<br>months |
| 17)<br>469<br>8 nodes | TBI | left MTS | normal | L MT | L ATL | gliosis<br>hippocampal<br>sclerosis | Engel IIIA@63<br>months |
| 18)<br>IO21 | None | Prior R. ATL | N/A | Right orbitofrontal<br>cortex. | R. Frontal lobe | hippocampal<br>sclerosis,<br>cortical<br>dysplasia | Engel<br>IVB@24<br>months |
| 19)<br>4110<br>10 nodes | encephalitis | Encephalomalacia | Normal | L inferior frontal<br>gyrus, insula, MT | L temporal lobe<br>and insula<br>resection | Gliosis | SUDEP<br>@6 weeks |
| 20)<br>IO23<br>20 nodes | Significant<br>head<br>injury<br>with<br>LOC | Left temporal T2<br>hyperintensity<br>with mild<br>enhancement | N/A | Bilateral MT,<br>right lateral<br>temporal | L temporal<br>lobectomy,<br>anterior thalamic<br>DBS | Gliosis | Engel<br>IVB@24<br>months |
| 21)<br>IO13<br>10<br>nodes | None | R parietal lobe<br>resection | R parietal and<br>R occipital | R insula, pre-<br>cuneus, middle<br>occipital gyrus,<br>superior parietal<br>lobule, superior<br>occipital gyrus,<br>superior temporal<br>gyrus, middle<br>temporal gyrus | R parietal | gliosis | Engel IIIA@18<br>months |
| 22)<br>IO15 | None | L posterior fossa<br>arachnoid cyst, R<br>ATL | R<br>temporal | L MT, R cingulate,<br>post. cingulate,<br>mesial frontal,<br>precuneus | R anterior<br>cingulate thermal<br>ablation | gliosis | Engel<br>IVB@36<br>months |
| 23)<br>IO19 | None | Prior R parietal<br>resection | R parietal<br>and<br>occipital<br>hypometabolism | R parietal lobe | R. Parietal lobe<br>resection | Gliosis | Engel<br>IVB@36<br>months |

Two patients who had resections (4122 and 479) had low FR rates and no SEEG contacts recorded fast ripples on spikes (fRonS) at a rate greater than 1 per minute. Since fRonS are believed to be a biomarker of epileptogenic tissue^13,15,36,50^, we concluded these patient’s SEEG implant had poor spatial sampling of epileptogenic regions (Table 1, Figure 1). Patient 4122 had a seizure free outcome after a modified right anterior temporal lobectomy (ATL), but patient 479 had an Engel IV outcome after a modified right ATL.

Another three patients (456, 473, and IO021) not in the resection cohort had nearly all FR sites removed, but were not seizure free (Table 1, Figure 1). Patient 456 had bilateral temporal lobe seizures and underwent an ATL with an Engel IV outcome. Patient 473 suffered a traumatic brain injury, had widespread seizure onsets, and underwent a thermal ablation of the left mesial temporal region. Lastly, patient IO021 had a right ATL but continued to have widespread seizure onsets and later underwent a second resection of the right frontal lobe (Figure 1, Table 1). In these patients, we assumed the spatial sampling of epileptogenic regions by the SEEG contacts was incomplete. Since our study was retrospective these patients could be excluded from the study cohort of 18 patients (Table 1, Figure 1, see discussion).

A second cohort of 10 patients with RNS included seven males and three females between the ages of 29 and 58 years old. Three of the ten patients treated with RNS were classified as super responders (>90% reduction in seizure frequency) and the remaining seven as intermediate responders^59^(Figure 1, Table 2). No patient was classified as a poor responder. Outcome was assessed 4 years or longer after RNS surgery. (Table 2). None of these patients met the criteria for poor spatial sampling.

**Table 2:**
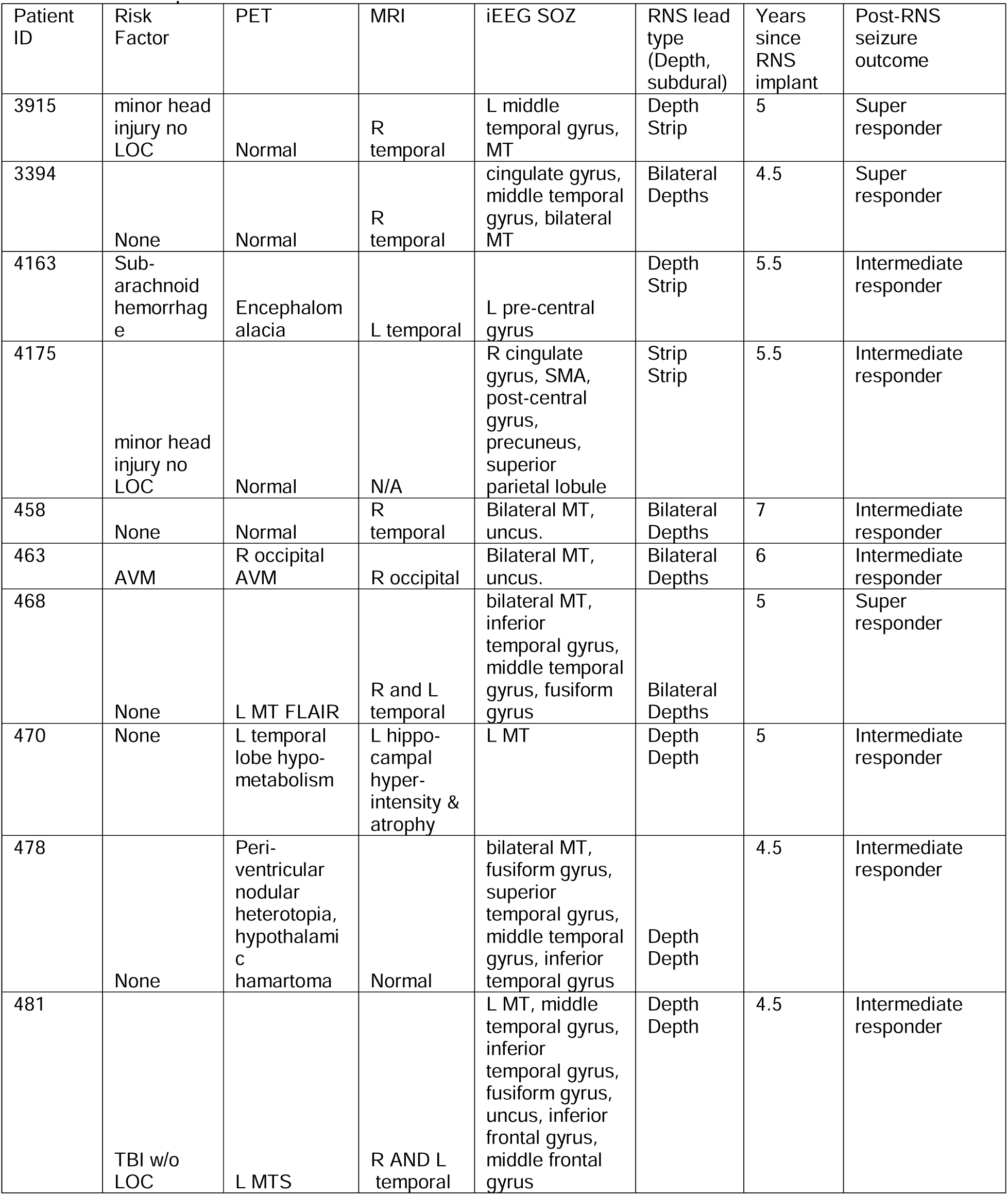
Responsive neurostimulator system (RNS) patient characteristics. Note RNS lead type is not bilateral unless specified. Abbreviations: L: left, R: right, LOC: loss of consciousness, AVM: arteriovenous malformation, MT: mesial temporal

| Patient ID | Risk Factor | PET | MRI | iEEG SOZ | RNS lead type (Depth, subdural) | Years since RNS implant | Post-RNS seizure outcome |
| --- | --- | --- | --- | --- | --- | --- | --- |
| 3915 | minor head injury no LOC | Normal | R temporal | L middle temporal gyrus, MT | Depth Strip | 5 | Super responder |
| 3394 | None | Normal | R temporal | cingulate gyrus, middle temporal gyrus, bilateral MT | Bilateral Depths | 4.5 | Super responder |
| 4163 | Sub-arachnoid hemorrhage | Encephalomalacia | L temporal | L pre-central gyrus | Depth Strip | 5.5 | Intermediate responder |
| 4175 | minor head injury no LOC | Normal | N/A | R cingulate gyrus, SMA, post-central gyrus, precuneus, superior parietal lobule | Strip Strip | 5.5 | Intermediate responder |
| 458 | None | Normal | R temporal | Bilateral MT, uncus. | Bilateral Depths | 7 | Intermediate responder |
| 463 | AVM | R occipital AVM | R occipital | Bilateral MT, uncus. | Bilateral Depths | 6 | Intermediate responder |
| 468 | None | L MT FLAIR | R and L temporal | bilateral MT, inferior temporal gyrus, middle temporal gyrus, fusiform gyrus | Bilateral Depths | 5 | Super responder |
| 470 | None | L temporal lobe hypometabolism | L hippocampal hyperintensity & atrophy | L MT | Depth Depth | 5 | Intermediate responder |
| 478 | None | Peri-ventricular nodular heterotopia, hypothalamic hamartoma | Normal | bilateral MT, fusiform gyrus, superior temporal gyrus, middle temporal gyrus, inferior temporal gyrus | Depth Depth | 4.5 | Intermediate responder |
| 481 | TBI w/o LOC | L MTS | R AND L temporal | L MT, middle temporal gyrus, inferior temporal gyrus, fusiform gyrus, uncus, inferior frontal gyrus, middle frontal gyrus | Depth Depth | 4.5 | Intermediate responder |

### Characterizing FR metrics

In the resection cohort of 18 patients, we compared the RR of: 1) FR>350 Hz (fRonO > 350 Hz and all fRonS); 2) all FR (200-600 Hz); 3) RonS (80-200 Hz); and 4) RonO (80-200 Hz) between the nine seizure free and nine non-seizure free patients. We found that only FR>350 Hz trended toward a higher RR in the seizure free than non-seizure free patients (Figure 2A, Kruskal-Wallis Chi-sq=2.13, p=0.15, n=18). The other HFO subtypes showed no trend or significant differences (Figure 2B-D).

**Figure 2:**
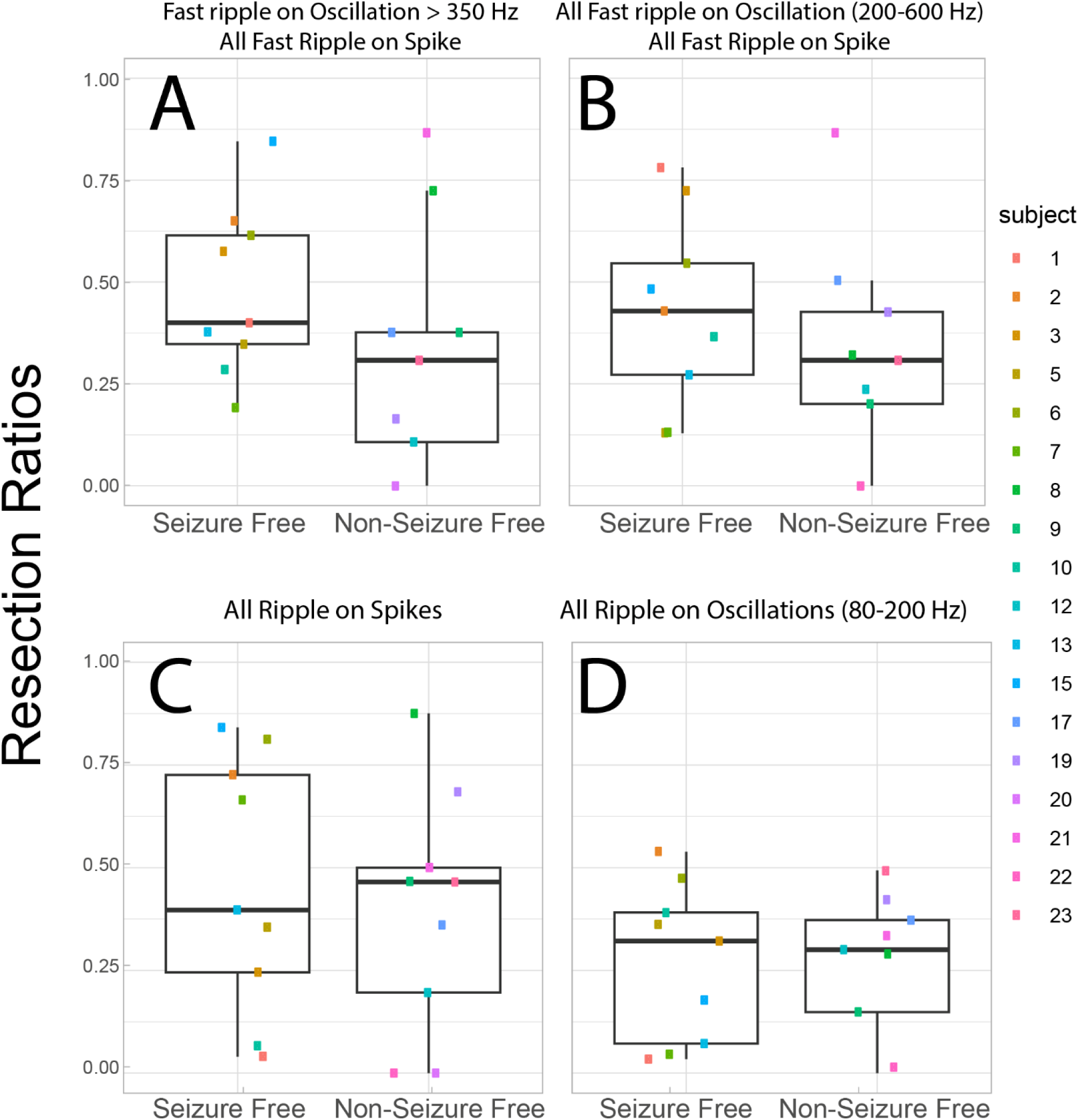
The resection ratio (RR) of higher frequency fast ripples (FR) better differentiates seizure free patients. (A) The RR of FR on oscillations > 350 Hz and all FR on spikes (200-600 Hz) trended higher in the seizure free than non-seizure free patients (Kruskal-Wallis Chi-sq=2.13, p=0.15, n=18). The other HFO subtypes including all FR (200-600 Hz, panel B), ripples on spikes (C), and ripples on oscillations (D) showed no trends or significant differences in the HFO RR (Kruskal-Walis p>0.2, n=18). Patient identification numbers labeled as in Table 1.

We then evaluated whether the FR graph theoretical measures derived from FR>350Hz would differ with respect to seizure outcome. The spatial FR net metric was significantly higher in the non-seizure free than seizure free patients (Figure 3B, Kruskal-Wallis Chi-Sq = 9.92, p=0.002, n=18). By contrast, the temporal FR net-A and temporal FR net-B metrics trended higher in the seizure free than non-seizure free patients (Figures 3C & 3D; Kruskal-Wallis Chi-Sq = 3.29, p=0.07, n=18 & Kruskal-Wallis Chi-Sq = 2.67, p=0.10, n=18). There was no obvious correlation between the four metrics, suggesting interdependence within and across patients (Figure 3).

**Figure 3:**
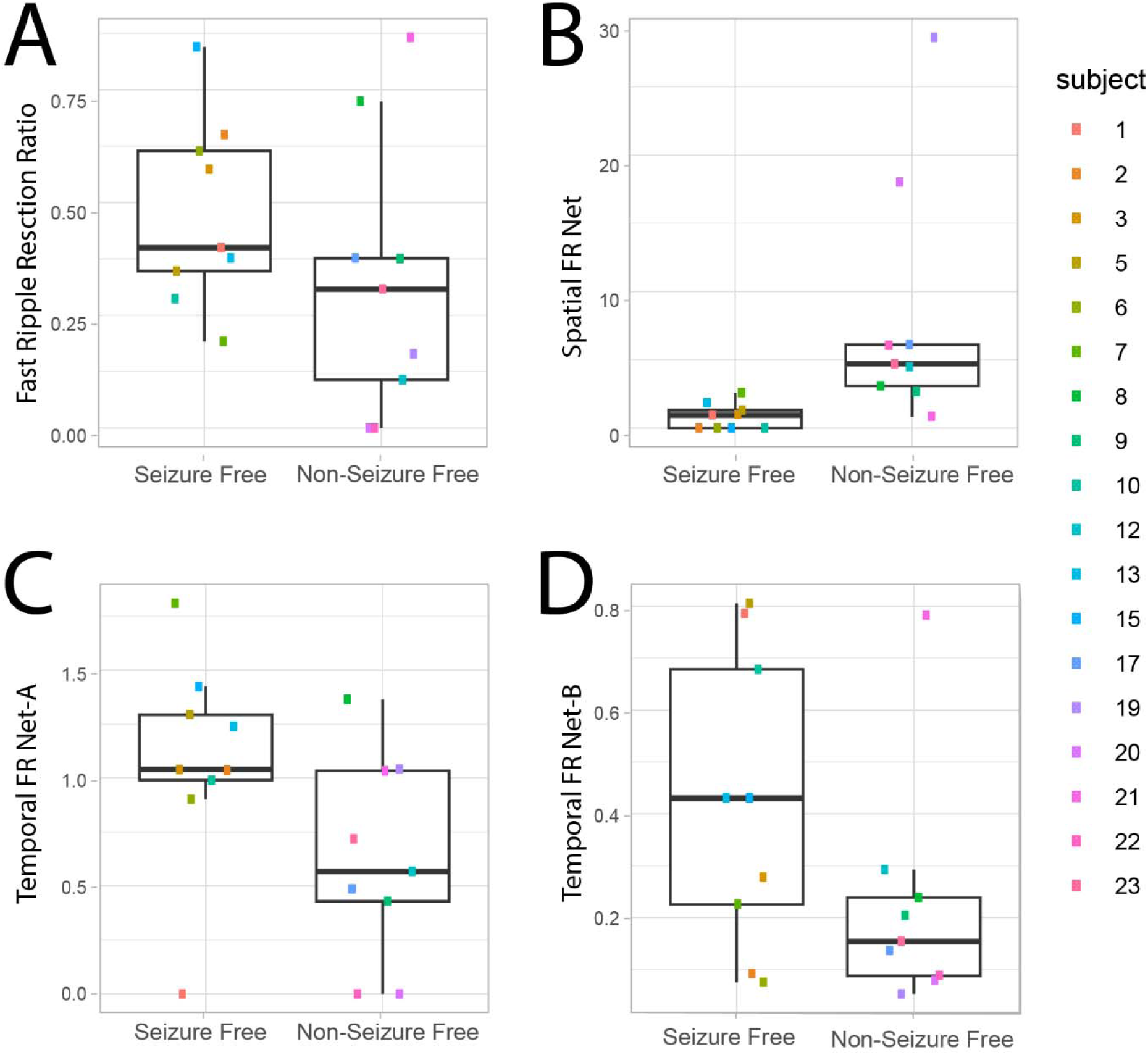
Four fast ripple (FR) factors used for training and testing a support vector machine (SVM) to label seizure free patients show differences in seizure free patients. The four factors used fast ripple (FR) on oscillation >350 Hz and all FR on spikes (200-600 Hz). (A-D) Box and scatter plots of the four metrics between seizure free and non-seizure free outcome. (A) The FR RR trended higher in seizure free patients (Kruskal-Walis Chi-sq=2.13, p=0.15). The spatial FR net metric was significantly higher in the non-seizure free patients compared with seizure free patients (Kruskal-Wallis Chi-Sq = 9.92, p=0.002, n=18). The temporal FR net-A metric trended (C) higher in the seizure free patients (Kruskal-Wallis Chi-Sq = 3.29, p=0.07, n=18). The temporal FR net-B metric also trended higher in the seizure free patients (Kruskal-Wallis Chi-Sq = 2.67, p=0.10, n=18). When all four of these metrics (FR RR, Spatial FR Net, Temporal FR Net-A, B) were used as factors to train a support vector machine to label seizure free patients, the SVM exhibited a 78% accuracy with leave one out cross-validation. Patient identification numbers labeled as in Table 1.

The four metrics (FR RR, Spatial FR Net, Temporal FR Net-A, B) were used as factors to train a SVM to label seizure free patients, and the SVM had a 78% accuracy with leave one out 18-fold cross-validation. We did not use other HFO subtypes to train the SVM because their RR performed poorly in distinguishing seizure free from non-seizure free patients (Figure 2B-D). Also, our past work showed aspects of the spatial and temporal HFO network measures perform sub-optimally when applied to other HFO subtypes like RonS or RonO^42,43^.

### Rationale for the virtual resection method targeting autonomous, high-rate FR nodes

Retrospective analysis of FR resection and post-operative outcome has used the FR RR. Two shortcomings of the FR RR are it does not specify what portion of FR need be resected to achieve a seizure free outcome in a prospective context, and second it poorly handles spatial sampling limitations. While the latter issue can be addressed with spatial FR net^43^, the former issue is unresolved. Using a graph theoretical analysis of FR temporal correlations (i.e., mutual information [MI] between the onset times of FR from different electrode contacts), we found FR with a rate greater than 1 per minute had a lower nodal local efficiency (LE). Lower LE indicates greater autonomy in generating FR at one node (i.e., SEEG contacts) with respect to FR at other nodes (Figure 4A). These results also imply that nodes with lower nodal FR LE have relatively lower FR MI edges, and the lower FR MI edges correspond with greater FR rates in both the paired nodes connected by the edge^60^. For all patients in the resection cohort the number of total nodes in FR MI network with a nodal local efficiency greater than zero are shown in Table 1. Using k-means clustering to select autonomous, high-rate FR nodes (Figure 4A, blue cluster) we found a significantly greater number of unresected autonomous, high-rate nodes in not seizure free than seizure free patients (Figure 4B, Kruskal-Wallis Chi-sq 5.62, p=0.02, n=18). This result supports the utilization of temporal FR net-A, B measures for classifying seizure freedom in patients, and targeting autonomous, high-rate FR nodes in a virtual resection.

**Figure 4:**
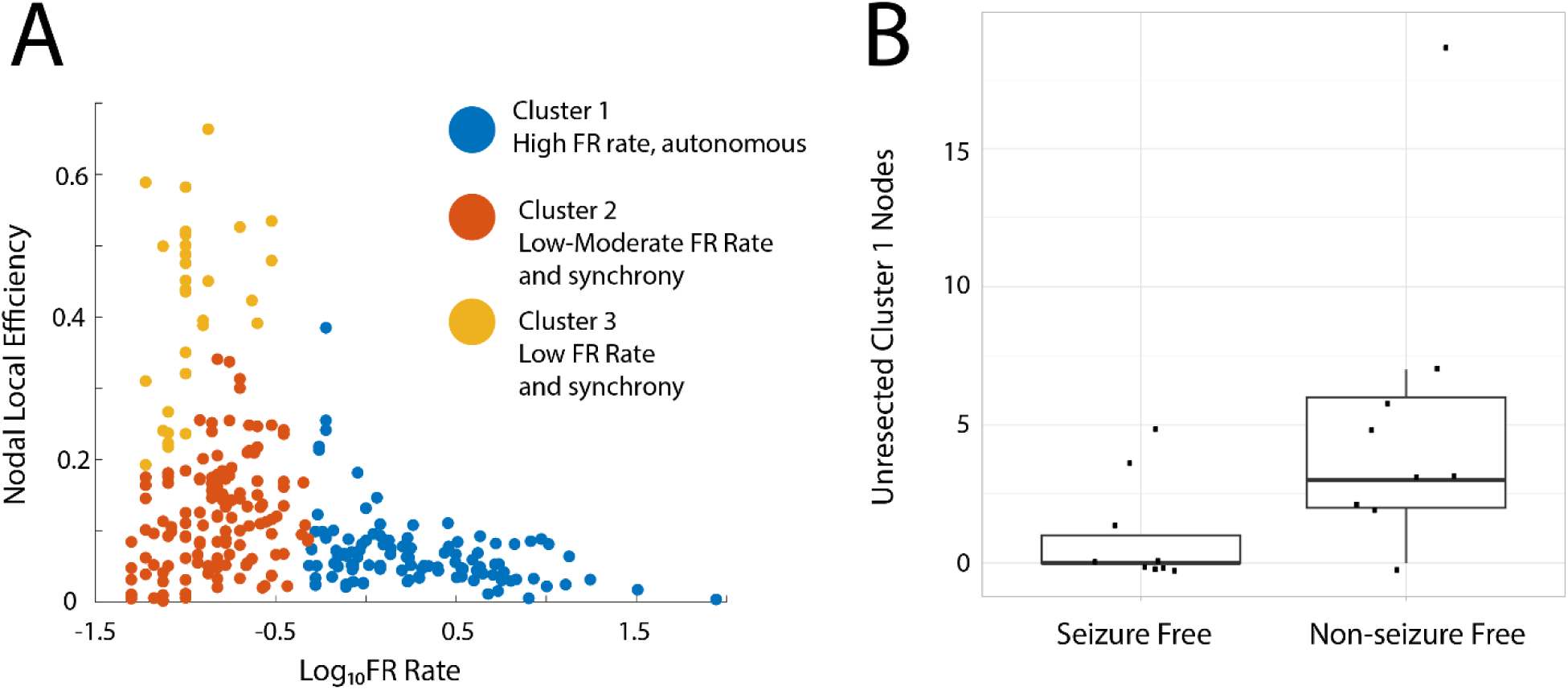
A failure to resect tissue proximal to electrode contacts generating autonomous, high-rate fast ripples (FR) sites correlate with a non-seizure free outcome. (A) K-means clustered scatter plot of the logarithm of the FR rate (FR on oscillations > 350 Hz, and all FR on spikes) on the x-axis, and the corresponding nodal local efficiency on the y-axis. In cluster #1 (blue) the electrode contacts (i.e., nodes) higher rates of FR had a lower nodal local efficiency. Low local efficiency corresponds with lower mutual information and more autonomy in FR generation (i.e., a loss of synchrony). (B) Box and scatter plot of the number of Cluster #1 nodes (unresected; points in blue in panel A) in each of the 18 patients dichotomized as seizure free and non-seizure free. Patients with non-seizure free outcomes had a significantly larger number of unresected autonomous, high-rate FR sites (Kruskal-Wallis Chi-sq 5.62, p=0.02, n=18).

**Figure 5:**
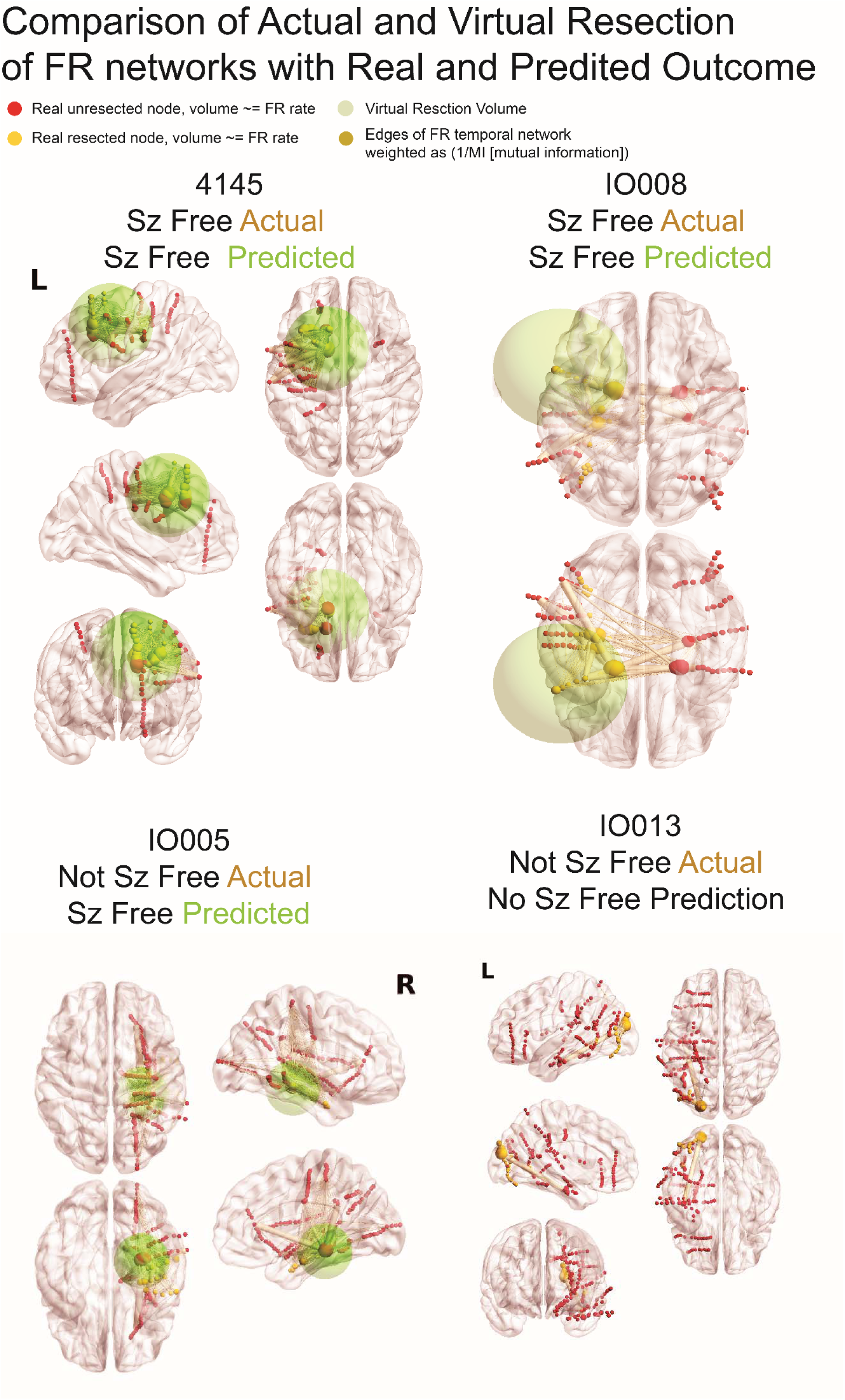
Illustration of fast ripple (FR) networks and real and virtual resections. In the four patients, the sizes of red (unresected) and yellow (resected) nodes [i.e., stereo-EEG electrode contacts] are proportional to the relative FR rate. The edges (pale yellow), connecting the nodes to one another, are weighted in size by the inverse of the mutual information (MI) of FR temporal correlations between the two nodes. The green sphere denotes the borders of the virtual resection. The center of the sphere is the node with most FR autonomy and/or highest FR rate and has a margin of 1 cm. The four FR metrics (FR resection ratio, spatial FR net, and temporal FR net-A, B) are derived from comparison of the sets of FR generating contacts in and outside of the virtual resection sphere and these factors are used by the support vector machine (SVM) to predict virtual seizure (sz) freedom. If the SVM predicts non-seizure (non-sz) freedom the virtual resection model iterates, and the virtual resection sphere may expand depending on whether the spatial location of the next top node that generates FR at higher rates and most autonomy is outside the sphere. In the case that the sphere expands, the new margins are extended by 1 cm. If the virtual resection includes three lobes it is considered a failure. As shown, extension of the virtual resection sphere outside spatially sampled regions, and outside the brain, does not increase the number of nodes in the virtual resection set. Contralateral nodes within the virtual resection sphere are also excluded from the virtual resection set of nodes. For patients 4145 and IO008, who were rendered seizure free, the virtual resection that was predicted as sufficient for virtual seizure freedom, included a set of nodes that partially overlaps with the set of resected nodes. In patient 4145 the contacts in the virtually resected set (red and yellow nodes within green sphere) were larger than the set of resected nodes (yellow nodes). In patient IO008 the difference between the set of nodes in the virtual resection and actual resection was smaller (see table 3). Patient IO005 was not seizure free, but the virtual resection that predicted a seizure free outcome was more posterior and included nodes with high FR rate and low MI edges. Patient IO013 was not seizure free, but the virtual resection did not produce seizure freedom because it required a resection of the occipital, parietal, and temporal lobe.

### Comparison of the volume and overlap of actual resections and FR virtual resections

In the resection cohort, a virtual resection was performed in each individual patient. In brief, the virtual resection methodology (see Methods) consisted of an iterative process where the first iteration selected the node with greatest autonomy (i.e., lowest nodal LE) and/or highest FR rate as the center of a sphere with a 1 cm radius representing the resection. Then the four metrics using FR>350 Hz metrics (see “Characterizing FR metrics”) were computed using nodes within and outside the sphere. The metric values were then used as factors in the trained SVM to classify the virtual resection as seizure free or not seizure free. If the classification was not seizure free and the next candidate autonomous FR node was outside the original sphere, a second iteration was performed where the sphere was expanded to include the next candidate FR node with a 1 cm margin. The four FR metrics were recalculated using nodes within and outside the revised sphere and then tested using the SVM. This process was repeated until classification was seizure free or the resection extended into three lobes.

Using virtual resections targeting autonomous, high rate FR sites we found all nine of the seizure free patients could be virtually seizure free. In all but one patient, the radius of the virtual resection was larger than the actual resection (Wilcoxon signrank, p=0.04, n=9) (Figure 5,6, Table 3). Comparing the set of contacts between the virtual and actual resection in the seizure free patients showed a mean accuracy of 0.63+/−0.06 and an F1 score of 0.50 +/− 0.07 (Table 5). Despite the lack of agreement, the set of electrode contacts in the virtual resection had a SOZ RR of 0.88+/−0.06 and a RonS RR of 0.76+/−0.08 (Table 3). We also examined the trends in the four FR metrics at different iterations of the virtual resection. We found that incrementally larger spheres increased the FR RR, decreased spatial FR net, and increased the FR spatial net-B. However, the temporal FR net-A could paradoxically decrease (Figure 6). This unexpected result is due to small resections that target the most autonomous FR site, which increased the numerator of the temporal FR net-A value. This is contrary to our results in Figure 3C where higher FR net-A values correlated with seizure freedom (Figure 3C).

**Figure 6:**
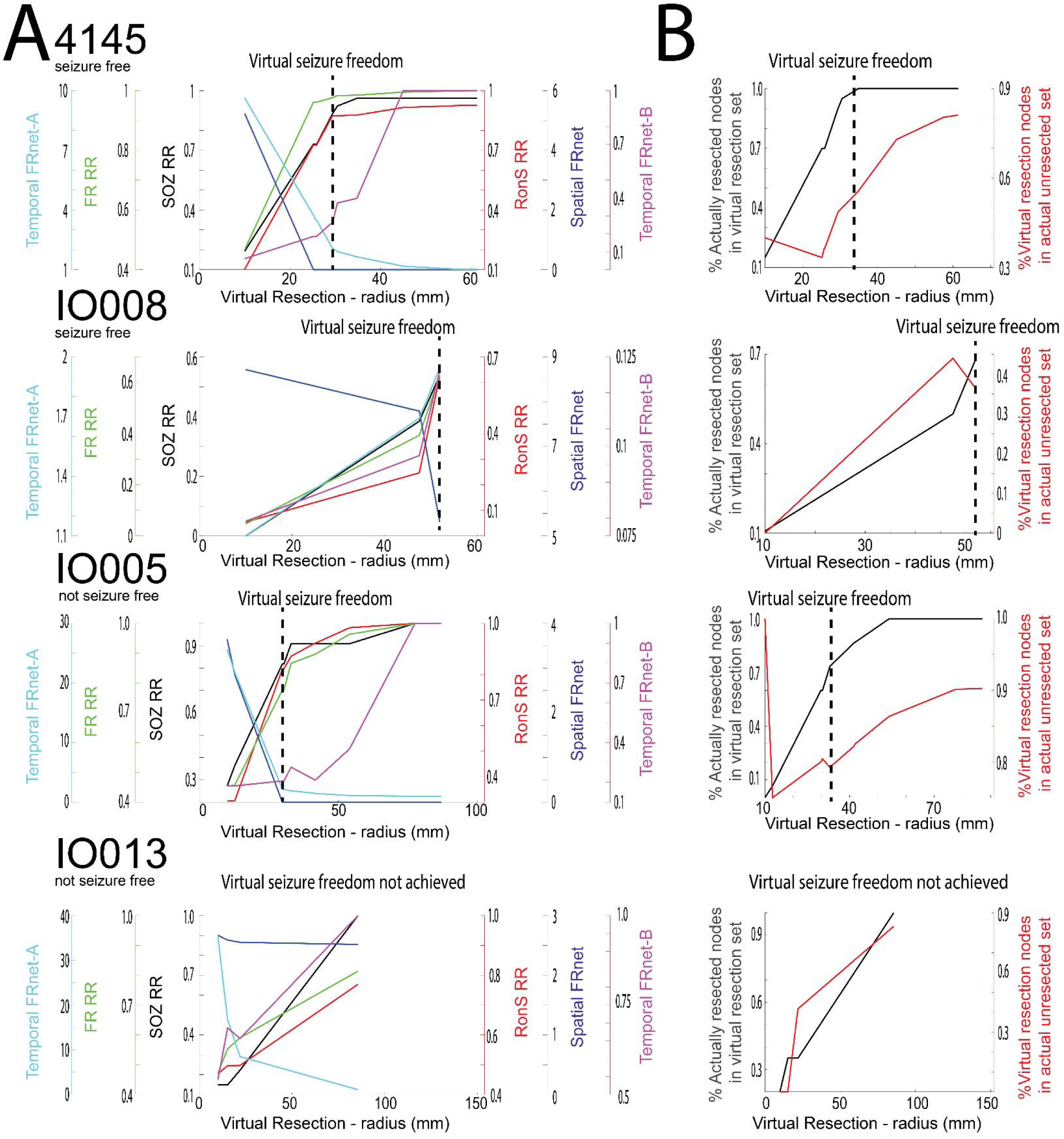
Changes in resection metrics in individual patients at different resection volumes. A) Comparison of the seizure onset zone resection ratio (SOZ, RR), ripple on spike RR (RonS RR, red), fast ripple RR (FR RR, green), spatial FRnet (blue), temporal FRnet-A (cyan), temporal FRnet-B (magenta) for two seizure free example patients (top), and two non-seizure free example patients (bottom). The hashed vertical line denotes the virtual resection iteration at which virtual seizure freedom is first achieved. Among the four patients, only in IO013 the virtual resection did not produce a seizure free outcome. B) Corresponding plot of the percentage of resected nodes in the resection set (black), and percentage of the virtual resection set in actual unresected nodes (red) for each of the four patients.

**Table 3:**
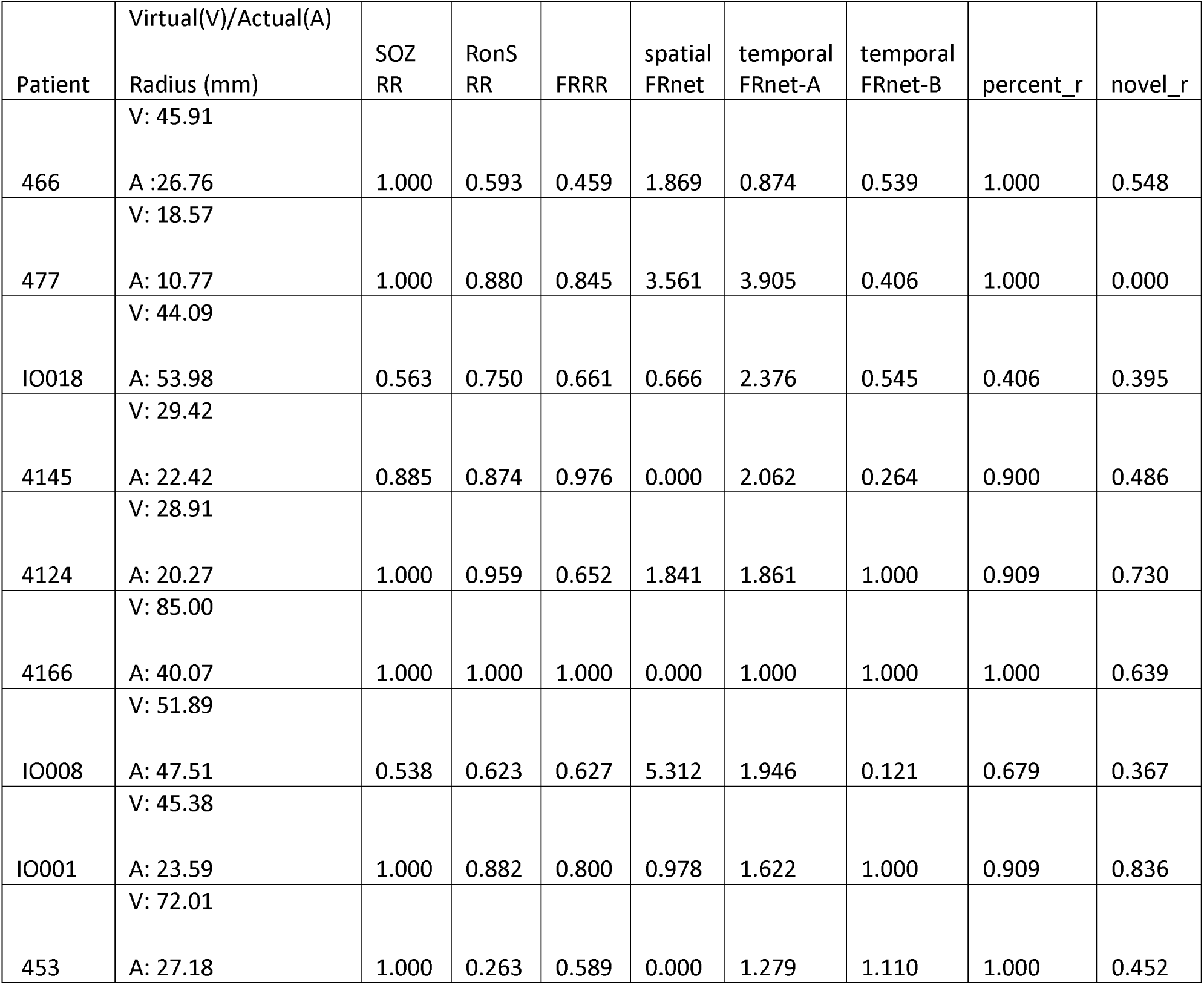
Metrics in patients who were seizure free and predicted to be seizure free with the virtual resection. Abbreviations: SOZ: seizure onset zone; RR: resection ratio; FR; fast ripple: FRnet: FR network graph theoretical measure; percent_r: percent of the nodes (i.e., electrode contacts) in resection cavity included in virtual resection set; novel_r: percent of nodes in the resection set that were not in the resection cavity; V: virtual radius of resection in mm; A: actual radius of resection in mm. In these patients who were seizure free, the virtual resection had a larger radius than the actual resection (Wilcoxon signrank, p=0.04, n=9).

We next computed virtual resections in patients who were not seizure free. We found that virtual resections targeting autonomous, high rate FR nodes could achieve seizure freedom in 5 of 9 of subjects (Table 4). In the virtually seizure free patients, excluding IO023 who had a small resection of cortex not sampled by the SEEG implant, the virtual resection radius was larger than the actual resection radius (Table 4, Figure 5,6 Wilcoxon signrank, p=0.02, n=4). Comparing the set of nodes between the virtual and actual resection, the four patients had a mean accuracy of 0.66 +/− 0.09 and F1 score of 0.45 +/− 0.102 (Table 6), which was similar to the nine seizure free patients. In 3 of the 4 patients, the virtual resection included nodes in the SOZ and high rates of RonS. The mean SOZ RR was 0.68+/−0.17 and the RonS RR was 0.78+/−0.16 (Table 4). Only patient 4110 had poor overlap of nodes between the virtual and actual resection. The study wasn’t adequately powered to compare accuracy and F1 score of the virtual resection between seizure free and not seizure free patients.

**Table 4:** Metrics in patients not seizure free and who were predicted to be seizure free by the virtual resection. Four patients did not achieve seizure freedom from the virtual resection (red text). Abbreviations: SOZ: seizure onset zone; sim: simulation; RR: resection ratio; FR; fast ripple: FRnet: FR network graph theoretical measure; percent_r: percent of the nodes (electrode contacts) in resection cavity included in virtual resection set; novel_r: percent of nodes in the resection set that were not in the resection cavity. V: radius of virtual resection in mm; A: radius of actual resection in mm. In the five patients who were predicted to be seizure free from the virtual resection, the resection radius trended larger than the actual resection (Wilcoxon signrank, p=0.02, n=4).

| Patient | Virtual(V)/Actual(A)<br>Radius (mm) | SOZ<br>RR | RonS<br>RR | FRRR | spatial<br>FRnet | temporal<br>FRnet-A | temporal<br>FRnet-B | percent_r | novel_r |
| --- | --- | --- | --- | --- | --- | --- | --- | --- | --- |
| 10)<br>IO005 | V:29.85<br>A:22.40 | 0.818 | 0.821 | 0.776 | 0.000 | 2.222 | 0.207 | 0.600 | 0.800 |
| 11)<br>IO012 | A: 18.42 | 1.000 | 0.879 | 0.737 | 3.083 | 4.898 | 0.084 | 0.818 | 0.471 |
| 12)<br>469 | V:53.05<br>A:33.44 | 1.000 | 0.824 | 0.947 | 0.000 | 1.249 | 0.353 | 0.857 | 0.478 |
| 13)<br>4110 | V:35.08<br>A:27.11 | 0.318 | 0.191 | 0.833 | 0.000 | 1.282 | 0.088 | 0.162 | 0.905 |
| 14)<br>462 | V:75.66<br>A:26.06 | 1.000 | 0.906 | 0.429 | 4.360 | 0.894 | 0.262 | 1.000 | 0.600 |
| 15)<br>IO023 | V:74.12<br>A: 9.02 | 1.000 | 0.935 | 0.942 | 0.000 | 1.206 | 0.096 | 0.000 | 1.000 |
| 16)<br>IO013 | A: 85.60 | 1.000 | 0.769 | 0.814 | 2.517 | 1.076 | 1.000 | 1.000 | 0.832 |
| 17)<br>IO015 | A: 50.24 | 0.625 | 0.992 | 0.915 | 3.261 | 1.629 | 0.099 | 0.000 | 1.000 |
| 18)<br>IO019 | A: 64.90 | 1.000 | 0.998 | 0.932 | 2.602 | 1.001 | 1.000 | 1.000 | 0.589 |

**Table 5:**
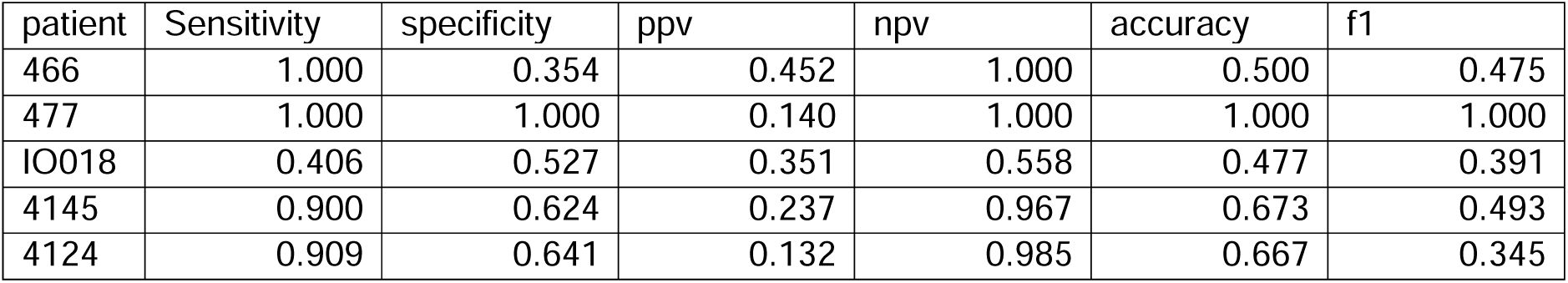
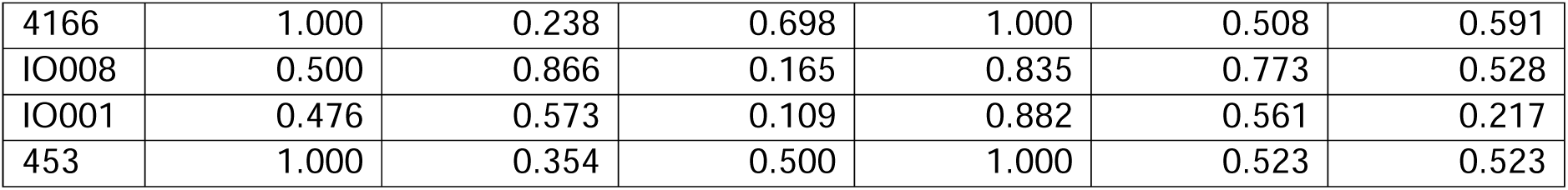
Sensitivity, specificity, positive predictive value (ppv), negative predictive value (NPV), accuracy and f1 score (F1) comparing the resected electrode contacts in patients seizure free with the resected contacts by virtual resection predicted to produce a seizure free outcome.

**Table 6:** Sensitivity, specificity, positive predictive value (ppv), negative predictive value (NPV), accuracy and f1 score (F1) comparing the resected electrode contacts in patients who were not seizure free with the resected contacts by virtual resection predicted to produce a seizure free outcome.

| patient | sensitivity | specificity | ppv | npv | accuracy | f1 |
| --- | --- | --- | --- | --- | --- | --- |
| IO005 | 0.600 | 0.928 | 0.065 | 0.956 | 0.896 | 0.529 |
| 469 | 0.857 | 0.324 | 0.522 | 0.846 | 0.479 | 0.490 |
| 4110 | 0.162 | 0.655 | 0.077 | 0.699 | 0.531 | 0.148 |
| 462 | 1.000 | 0.681 | 0.273 | 1.000 | 0.746 | 0.615 |

### Simulations of virtual placement of RNS at high-rate FR sites

For individual patients with RNS we used measures of proximity between the RNS stimulation contacts and the presurgical SEEG contacts to compute the SOZ stimulation ratio (SR), the FR SR, and a graph theoretical measure the RNS temporal FR net. In our cohort of 10 patients, we found that only in 3 patients who had a super response (>90% seizure reduction) trended towards a higher SOZ SR, a significantly increased FR SR, and a significantly decreased RNS temporal FR net^54^. The decreased RNS temporal FR net indicates the proximity of stimulating contacts to autonomous, high rate FR sites (Figure 7). Based on these preliminary findings we asked if virtual RNS stimulation contacts selected contiguous to the pre-surgical stereo EEG contacts with highest FR rate would result in a higher SOZ SR, FR SR, and lower RNS temporal FR net. We found that repositioning the stimulation contacts to these pre-surgical SEEG sites did not influence the values for super responders. However, one intermediate responder had a higher SOZ SR, higher FR SR, and lower RNS temporal FR net value, while another intermediate responder showed a lower SOZ SR, but increased FR SR and lower RNS temporal FR net value (Figure 7).

**Figure 7:**
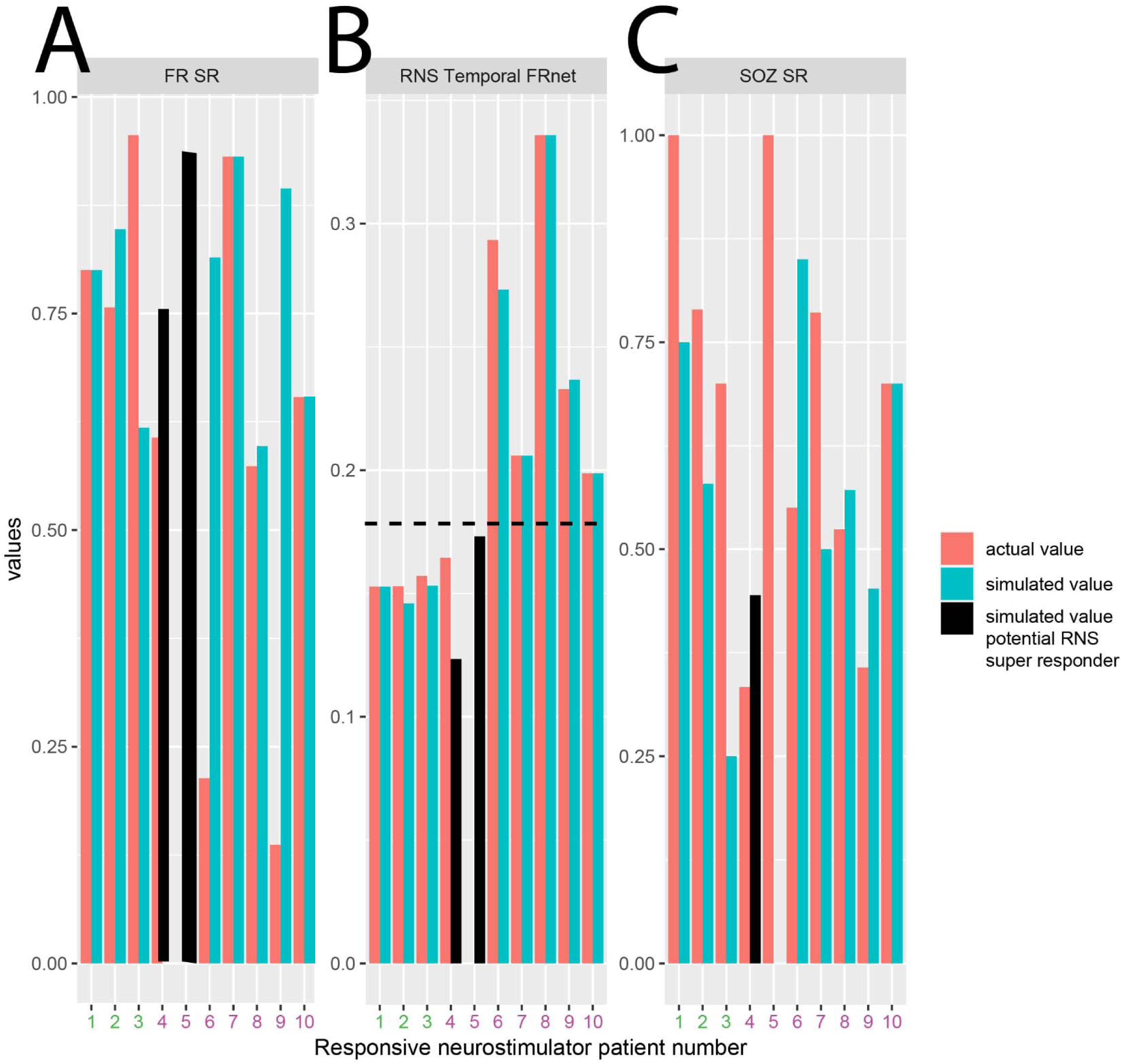
Simulated responsive neurostimulator lead placement and metrics that may predict RNS seizure outcome response. The actual fast ripple stimulation ratio (A, FR SR), RNS temporal FR net (B), and seizure onset zone stimulation ratio (C, SOZ SR) are shown as orange bars for the ten patients in the RNS cohort. Patients 1-3 (patient id#: 3915,3394,468, see Table 2) had a clinical super response (>90% seizure reduction). Patients 4-10 had an intermediate responder outcome. The actual RNS temporal FR net values were significantly lower in patients 1-3 compared to patients 4-10 (Kruskal-Wallis Chi-sq 5.4, p=0.02, n=18). The simulated FR SR, RNS temporal FRnet, and SOZ SR, shown in cyan bars, are derived from superposition of the virtual RNS stimulation contacts to contiguous pre-surgical stereo EEG contacts with highest FR rate. In patients 4-5 (patient id#: 478, 4163, black bars, see Table 2) the RNS temporal FR net decreased (B, horizontal hatched line), and the FR SR increased (A). In patient 4, but not in patient 5, the virtual stimulation contacts were more proximal to the SOZ than the actual stimulation contacts (C). This simulation suggests that measuring the SOZ SR, FR SR, RNS temporal FRnet associated with virtual RNS placement may increase the odds of super responder outcome.

## Discussion

In this retrospective study, we trained a SVM using four FR metrics and seizure free, or not, seizure outcome labels. These metrics are derived from differences in the rate, spatial distance, and temporal interdependencies of FR within and between SEEG contacts with respect to the actual resection cavity. The rationale for utilizing the temporal FR net-A and B measures in the SVM and targeting high rate autonomous cortical FR sites for virtual resection was that a failure to resect such FR sites correlated with non-seizure free outcome. Leave one out cross-validation showed that the SVM trained on the four FR factors, in the 18 patients, performed with 78% accuracy like the results of Nevalainen et al.^12^. To test the SVM in our study, we didn’t have a separate test cohort; rather, the SVM labeled seizure free outcome, or not, in each simulated iteration of the virtual resections for the same 18 patients. We found that in all but one surgical case, that were performed using the standard of care and sufficient for seizure freedom, the actual resection was smaller than the virtual resection labeled as seizure-free. One explanation is resection guided by the standard of care use multiple modalities, especially neuroimaging. In contrast, the virtual resections solely relies on inter-ictal FR using arbitrary ratios and cut-off values^61^. In support of this explanation, in patients rendered seizure free, the virtual resection showed greater than 75-85% overlap with the SOZ and RonS sites^13^, but an f1 score of 0.5 with the actual resection. Alternatively, the standard of care-based resections can be restricted by eloquent cortical regions. SVM-based virtual resections could still play a significant role in improving epilepsy surgery outcomes. Assuming the SVM accurately classifies seizure free outcome, then prospectively patients predicted to fail the standard of care resection could undergo simulated virtual resections. The SVM virtual resection removing FR associated with a seizure free outcome could amend the standard of care resection and thereby increase the odds of seizure freedom^61^.

### Power calculations for use of virtual resections in a randomized controlled clinical trial

The ultimate goals in the surgical and RNS treatment of medication-resistant epilepsy are the elimination or complete control of seizures with minimal morbidity. Towards these goals results from our simulations could be used in power calculations to design a randomized controlled trial (RCT) to compare two approaches to epilepsy surgery, i.e., a control arm using the clinical standard of care to guide resection, and an active arm that considers results from the SVM model to inform the surgical resection. The active arm would use the SVM model to predict whether a standard of care resection produces a seizure free outcome and in the event it doesn’t, then the virtual resection targeting sites important for the FR RR, spatial– and temporal-FRnet could be used to amend the original surgical plan. The decision to continue with the modified resection plan would be contingent on the agreement from the patient and the neurosurgeon.

A power analysis using our SVM model indicates a sample size of 150 patients in each arm will provide 80% power to detect a difference of 0.15 in seizure freedom rate between the control and active arms. This assumes the control arm has 60% seizure freedom and the active arm, benefitting from SVM-guided amendments, a significantly higher seizure freedom of 75%. The difference in seizure freedom between the approaches is consistent with our preliminary results showing an SVM classification accuracy near 0.8 and virtual resections based on spatial– and temporal FRnet measures that predicted a seizure free outcome in five of nine subjects. Two-sided Z-test with unpooled variance was used at a significance level of 0.05, to rigorously evaluate the efficacy of incorporating FR net analyses into surgical planning for epilepsy treatment. Anticipating a differential dropout rate because of 1) an inability to fully resect the SOZ due to overlap with eloquent cortex; 2) the refusal by patients and or physician for amended resections; and 3) patients who are lost to follow up with approximately 25% expected in the active arm compared 10% in the control arm. To account for these participant losses the RCT would need to enroll 200 and 167 subjects in the active arm and control arm, respectively, to maintain the power to detect differences in seizure freedom.

### Approaches for predicting and modeling seizure outcome in patients with RNS

RNS therapy was initially thought to reduce seizure frequency by detecting seizures and stimulating to abort the seizure^62^. However, the RNS device stimulates the brain over 1000 times a day and almost entirely during the inter-ictal epoch^63^. Seizure frequency decreases gradually over years following RNS^64^. Furthermore, closed– and open-loop stimulation have been shown to similarly effective^65^. One study found that a reduction in seizure frequency with RNS therapy correlated with an increased coherence in the low frequency intracranial EEG between 1 and 3 years post implant^59^. Thus, the efficacy of RNS may be more strongly related to induced alterations in the epileptic network^59,66^, and FR are critical nodes in this network. This explanation is consistent with the current modelling results that showed stimulating the highest rate FR sites reduced the size of the RNS temporal FRnet in the three super-responder patients and two intermediate responder patients (Figure 3). This suggests that targeting RNS stimulation to highly active FR sites could improve seizure control with RNS.

Our small pilot study of RNS responders lacked the power to use an SVM. Ideally in a larger study, the SOZ SR, FR SR, and RNS temporal FR net would be used together as factors to train and test an SVM to label RNS super responders. Should these experiments succeed, a larger cohort that could then be used to plan a prospective clinical trial. Based on current results, enrolling a total of 20 patients, divided equally into two groups: a control arm with standard of care RNS placement and an active arm using SVM results to inform the standard RNS placement, would achieve 80% power to detect a difference 0.5 between the group proportions of super responders. The proportion of super responders in the control arm is assumed to be 0.3. The test statistic used is the two-sided Z-Test with unpooled variance at 0.05 significance level. A critical goal of this aim is to accurately estimate the effect size of our intervention, which is pivotal for the planning of future, more extensive research.

### Alternative strategies for virtual resections and SVM training and testing

The current work focused on specific sub-population of FR>350 Hz. These FR were selected based on our prior studies using this cohort of patients showing that the FR>350 Hz were increased in the SOZ and resection margins of seizure free patients^42,43^. Other studies have shown that higher frequency FR may be more specific for epileptogenic regions^67,68^. Moreover, in murine models of epileptogenesis, FR>350 Hz are thought to signify greater importance in seizure genesis and may be generated by distinct mechanisms involving reduced spike-timing reliability^69,70^. Herein we found that the RR of other HFO subtypes like all FR, RonS, or RonO, were not different between seizure free and not seizure free patients, which contrasts with results from previous work^2,26,71^ and may be attributed to the unique clinical features of our study cohort. Nonetheless, in the current study, we did not derive spatial and temporal graph metrics with these HFO subtypes, but did perform this analysis in a prior study where we found these other HFO subtypes did not perform as well as FR>350 Hz in distinguishing better outcome patients^42^. In planned future studies with a larger sample size, we will assess the RR and spatial and temporal graph metrics for all the HFO types, including all FR (200-600 Hz).

Other HFO metrics can be used as factors to train diverse types of machine learning to label post-operative seizure outcome, and a SVM is just one of many types of machine learning that can be implemented (see methods). Opensource and collaborative efforts can help find the best method for using FR to predict outcome and plan virtual resections. For instance, our results show that the temporal FR net-A metric, which trended higher in seizure free patients, was paradoxically elevated in virtual surgeries labeled by the SVM as non-seizure free. This could be due to small resections targeting nodes with lowest LE and this increases the numerator of the temporal FR net-A metric defined as the RR of the FR MI path length. In future research, we will explore whether the unresected FR MI path length can be used as an alternative to the temporal FR net-A.

### Limitations

Like the study by Nevalainen et al.,^12^ we excluded patients with limited spatial sampling, which totaled 5 patients who were excluded from SVM training, cross-validation, and testing. Two of the 5 patients were excluded due to poor spatial sampling (*i.e.,* no FR MI network, no FR on spike > 1/min), and 3 patients were excluded due to incomplete spatial sampling (*i.e.,* poor post operative seizure outcome despite resection of the whole FR MI network). While patients with poor spatial sampling can be found prospectively using neurophysiological criteria, this is not the case for patients with incomplete spatial sampling because the post-operative seizure outcome is unestablished. One solution is finding patients with complete FR MI network resection in a large retrospective cohort and using these patient’s clinical, neuroimaging, neurophysiological data (excluding HFOs and FR) as factors, and the seizure free status as labels, to train a logistic regression model (LRM). Then in a prospective cohort, the trained LRM utilizes the same factors to predict a patient’s likelihood for a non-seizure free outcome (*i.e.,* incomplete spatial sampling). Another strategy to identify patients with incomplete spatial sampling is to investigate coupling of epileptic biomarkers with their spatial distribution and measuring the neurophysiological system’s response to coupling pertubation^72^.

Another limitation of this study was that the SVM was not trained and tested on distinct patients. However, the dangers of over training were minimized since the SVM was trained on actual resections then tested on virtual resections in the same patient cohort. Lastly our spherical resections may over-estimate the radius of the virtual resection cavity, thus unnecessarily including some SEEG contacts. More advanced geometric strategies to model the resection cavity may show smaller differences between the actual resection and virtual resections based on FR metrics.

Clinical, radiographic, and neurophysiologic factors, in the absence of inter-ictal HFO biomarkers, are also important in predicting post-operative seizure outcome^73,74^. We did not examine interactions between the trained SVM’s label and clinical factors in this study. Future work can use distinct LRMs that incorporate clinical, radiographic, and neurophysiologic factors and the label from the trained SVM. Understanding the interaction between the factors and the SVM label could be useful for defining the inclusion and exclusion criteria for a future clinical trial.

Lastly, we did not compare the FR-generating sites and the FR MI network with the neuroanatomic locations of lesions^75^. This comparison can be made in our future work to better understand whether the FR-generating tissue considered critical (i.e., autonomous, high-rate) overlaps with lesions such as focal cortical dysplasia.

### Conclusions

Our results indicate that autonomous, high-rate cortical sites generating FR>350Hz are most important for generating seizures. These FR sites can be used to predict if a resection defined by the standard of care will produce a seizure free outcome, and predict a seizure free outcome with a virtual resection that includes autonomous, high-rate FR sites. Virtual resections performed in this manner are larger in volume than the standard of care resection sufficient for seizure freedom. However, in cases where the standard of care resection is predicted to result in non-seizure freedom, amending the resection to include autonomous, high-rate FR sites could theoretically increase the odds of a seizure free outcome. Lastly, placing RNS to stimulate autonomous, high-rate FR sites may increase the odds of a super responder (>90% seizure reduction) outcome.

## Funding statement

This work was fully supported by the National Institute of Health K23 NS094633, a Junior Investigator Award from the American Epilepsy Society (S.A.W.), R01 NS106957(R.J.S.) and R01 NS033310 (J.E.).

## Author Contributions

Credit author statement: **Shennan Aibel Weiss:** conceptualization, methodology, software, investigation, resources, writing – original draft, writing – review & editing, funding acquisition. **Anli Liu**: investigation, writing – review & editing. **Werner Doyle:** investigation, writing – review & editing. **Charles Mikell III**: investigation, writing – review & editing. **Sima Mofakham**: investigation, writing – review & editing. **Noriko Salamon**: investigation, writing – review & editing. **Myung Shin Sim:** conceptualization, methodology, software, investigation, resources, writing – original draft, writing – review & editing, funding acquisition. **Itzhak Fried:** resources. **Chengyuan Wu:** resources. **Ashwini Sharan:** resources. **Jerome Engel:** writing – review & editing, funding acquisition. **Michael R. Sperling:** investigation, writing – review & editing, funding acquisition. **Anatol Bragin:** investigation, writing – review & editing. **Richard Staba:** investigation, writing – review & editing, funding acquisition.

## Data availability

The code for our fast ripple machine learning and virtual resections is published at https://github.com/shenweiss/frsvmsim. The MongoDB data for these simulations can be downloaded at https://zenodo.org/records/8125756.

## Disclosures Statement

M.R.S has received compensation for speaking at continuing medical education (CME) programs from Medscape, Projects for Knowledge, International Medical Press, and Eisai. He has consulted for Medtronic, Neurelis, and Johnson & Johnson. He has received research support from Eisai, Medtronic, Neurelis, SK Life Science, Takeda, Xenon, Cerevel, UCB Pharma, Janssen, and Engage Pharmaceuticals. He has received royalties from Oxford University Press and Cambridge University Press.

## Abbreviations

FR: fast ripple
fRonS: fast ripple on spike
fRonO: fast ripple on oscillation
MI: mutual information
LE: local efficiency
RNS: responsive neurostimulator
SEEG: stereo-EEG
iEEG: intracranial EEG
REM: rapid-eye movement sleep
SOZ: seizure onset zone

## Abbreviated summary

Weiss et al., show that utilizing FR metrics to plan an efficacious surgery is associated with larger resections than that of the standard of care. However, if FR metrics predict the standard of care resection will fail, the margins can be amended by certain FR sites to achieve seizure freedom.

## Acknowledgements

We would also like to thank Christina Louise George Trust and the Resnick Family Foundation.

## Notes

### Competing Interest Statement

S.A.W. has nothing to disclose, I.F. has nothing to disclose, C.W. Medtronic Inc. (advisory board), Micro Systems Engineering Inc. (advisory board), Neuropace Inc. (consultant), Nevro Corp. (consultant). J.E. has nothing to disclose. R.S. has nothing to disclose. M.R.S has received compensation for speaking at continuing medical education (CME) programs from Medscape, Projects for Knowledge, International Medical Press, and Eisai. He has consulted for Medtronic, Neurelis, and Johnson & Johnson. He has received research support from Eisai, Medtronic, Neurelis, SK Life Science, Takeda, Xenon, Cerevel, UCB Pharma, Janssen, and Engage Pharmaceuticals. He has received royalties from Oxford University Press and Cambridge University Press.

### Author Declarations

University of California Los Angeles Institutional Review Board Thomas Jefferson University Institutional Review Board

### Summary of Updates

This is an extensive revision of the original manuscript to meet the requirements of the reviewers.

